# Predicting Rectal Cancer Patient Survival with Dutch Radiology Reports using Natural Language Processing (NLP): The Role of Pretrained Language Models

**DOI:** 10.64898/2026.01.23.26344428

**Authors:** Lishan Cai, Tianyu Zhang, Regina Beets-Tan, Joren Brunekreef, Jonas Teuwen

**Affiliations:** Department of Radiology, The Netherlands Cancer Institute, Plesmanlaan 121, 1066 CX, Amsterdam, The Netherlands; GROW School for Oncology and Developmental Biology, Maastricht University Medical Centre, P. Debyelaan 25,66202 AZ, Maastricht, The Netherlands; Department of Medical Imaging, Radboud University Medical Centre, Geert Grooteplein 10, 6525 GA, Nijmegen, The Netherlands; Department of Radiation Oncology, Netherlands Cancer Institute, Plesmanlaan 121, 1066 CX Amsterdam, The Netherlands; Faculty of Science, University of Amsterdam, 1098 XH Amsterdam, The Netherlands

## Abstract

The use of Electronic Health Records (EHRs) has increased significantly in recent years. However, a substantial portion of the clinical data remains in unstructured text formats, especially in the context of radiology. This limits the application of EHRs for automated analysis in oncology research. Pretrained language models have been utilized to extract feature embeddings from these reports for downstream clinical applications, such as treatment response and survival prediction. However, a thorough investigation into which pretrained models produce the most effective features for rectal cancer survival prediction has not yet been done. This study explores the performance of five Dutch pretrained language models, including two publicly available models (RobBERT and MedRoBERTa.nl) and three developed in-house for the purpose of this study (RecRoBERT, BRecRoBERT, and BRec2RoBERT) with training on distinct Dutch-only corpora, in predicting overall survival and disease-free survival outcomes in rectal cancer patients. Our results showed that our in-house developed BRecRoBERT, a RoBERTa-based language model trained from scratch on a combination of Dutch breast and rectal cancer corpora, delivered the best predictive performance for both survival tasks, achieving a C-index of 0.65 (0.57, 0.73) for overall survival and 0.71 (0.64, 0.78) for disease-free survival. It outperformed models trained on general Dutch corpora (RobBERT) or Dutch hospital clinical notes (MedRoBERTa.nl). BRecRoBERT demonstrated the potential capability to predict survival in rectal cancer patients using Dutch radiology reports at diagnosis. This study highlights the value of pretrained language models that incorporate domain-specific knowledge for downstream clinical applications. Furthermore, it proves that utilizing data from related domains can improve the quality of feature embeddings for certain clinical tasks, particularly in situations where domain-specific data is scarce.

## Introduction

Colorectal cancer is the third most common cancer and the fourth leading cause of cancer-related mortality^1^. Rectal cancer accounts for about one-third of all colorectal cancer cases and its incidence keeps increasing^2^. An accurate survival prediction model for rectal cancer aids in personalized treatment, informed decision-making, and resource management, thereby improving patient outcomes and care quality. Several studies^3–6^ have applied imaging, clinicopathological, or molecular prognostic features to predict rectal cancer survival. However, these methods have shown limited effectiveness. Very few studies have focused on employing features derived from textual medical records for rectal cancer survival prediction. With the development of Electronic Health Record (EHR) systems, the volume of digital medical records has significantly increased^7^. However, around 80% of the records are unstructured without a standardized format and remain challenging to analyze for secondary use^8,9^. Manual feature extraction from EHRs is labor-intensive and introduces inter-reader variability. There is a high demand for accurate and robust models to extract features from unstructured medical records for downstream oncology research and clinical applications.

In the last decade, Artificial Intelligence (AI)-based rectal cancer research has gained increasing attention, especially in radiology and pathology^6,10,11^. AI has already demonstrated its value in the field of medical image analysis^12–15^. Beyond imaging, recent advancements in AI have showed significant capabilities in interpreting unstructured clinical texts and hold great potential for a range of downstream clinical tasks^16,17^. Several studies^18–22^ have demonstrated that deep-learning based Natural Language Processing (NLP) is capable of processing free-text medical reports to assist clinical decision-making. Many applications in cancer research — including disease progression, response to therapy, and survival outcomes — have utilized more specific medical records, such as radiology reports, which offer in-depth imaging findings, diagnostic interpretations, and treatment-related observations.

Radiology reports are often unstructured and complex, reflecting the varied terminology and descriptions used by different radiologists when documenting rectal cancer cases. To utilize language models for radiology reports, a widely adopted method^23^ involves fine-tuning the weights of a large language model to improve its representation for specific tasks. However, this approach requires a substantial amount of labeled data and significant computational resources^24^. The other common strategy for utilizing these reports is to use a pretrained language model to extract embeddings from the unstructured data for subsequent tasks. The advantage is that this decreases the need for labeled data specifically for training the task-specific large language model. However, this approach might lead to sub-optimal results if the extracted embeddings fail to capture informative nuances from the domain-specific free texts^25^. Kim et al.^20^ proposed a deep-transfer-learning model for predicting survival based on unstructured free-text data obtained from magnetic resonance imaging (MRI) reports of rectal cancer patients. In their study, feature vectors were extracted from MRI reports using Bidirectional Encoder Representations from Transformers (BERT) language models^26^. They compared several BERT variants, including Clinical BERT^27^, pretrained on publicly available clinical notes, BioBERT^28^, which is pretrained on large biomedical corpora, the original BERT^26^, and an un-pretrained BERT. Clinical BERT achieved the highest concordance index (C-index) of 0.595, although the differences between this and the other variants were negligible. The C-index, commonly used in survival analysis, measures the discriminatory ability of a model in ranking pairs of patients according to their predicted risk. A C-index of 1 indicates perfect concordance between predicted and actual outcomes, while a value of 0.5 suggests random performance. It is important to note that none of the pretrained models in their study had any specific knowledge of rectal cancer MRI reports. Verkijk et al.^29^ have shown that a domain-specific language model outperforming general models for domain-specific applications and when sufficient in-domain data is available, training a model from scratch appears to be more effective than extending the pretraining of an existing model, even if the latter includes a domain-specific vocabulary and retrains the embedding look-up layer. Similarly, Zhang et al.^21^ have pointed out that RoBERTa^30^, a robustly optimized BERT model, requires extensive pretraining data and publicly available models may not be suitable for specialized fields like medical tasks. They pretrained the model on a large dataset of Dutch breast cancer reports (in the millions) from scratch, enabling it to form a better understanding of their contents. The model then showed promising results in predicting the pathological outcomes of breast lesions through transfer learning.

In this study, we retrospectively collected Dutch EHRs (including radiology, pathology and endoscopy reports) from 1157 rectal cancer patients. Five RoBERTa models— including two publicly available (RobBERT and MedRoBERTa.nl) models and three developed in-house (RecRoBERT, BRecRoBERT and BRec2RoBERT) specifically for the purpose of this work — were compared for their ability to understand rectal cancer reports: RobBERT^31^, pretrained on the Dutch section of the OSCAR corpus; MedRoBERTa.nl^29^, which was trained using text data from Dutch hospital notes; RecRoBERT, pretrained solely on reports from 868 rectal cancer patients; BRecRoBERT, pretrained with reports from 868 rectal cancer patients and 37,517 breast cancer patients; and BRec2RoBERT, which involved continuing training BRecRoBERT using reports from 868 rectal cancer patients.

A key contribution of this study is the demonstration that domain-specific pretraining improves the representation of clinical reports, leading to improved performance in survival prediction tasks. We first evaluated the models on a masked word prediction task to assess their language understanding. Subsequently, we extracted feature representations from radiology reports at diagnosis and utilized them for downstream survival prediction. Our findings showed that BRecRoBERT, pretrained on both rectal and breast cancer datasets, produced the most informative embeddings, leading to superior predictions of overall survival (OS) and disease-free survival (DFS). These results highlight the value of domain adaptation in clinical NLP and suggest that integrating data from related medical domains can enhance model performance, particularly when direct task-specific data is limited.

## Results

### Patient Characteristics

In total, 15893 free-text medical reports including 3247 pathology (5.2 MB), 3244 endoscopy (5 MB) and 9402 radiology reports (13.9 MB), from 1157 patients in the Netherlands Cancer Institute (NKI) between January 2010 to December 2023 were retrospectively collected. All the reports were in Dutch. The total size of the corpus is 3,651,306 words. For all the radiology reports, the median word number of radiology reports is 219. Patients were included on the following criteria (1) biopsy-proven rectal adenocarcinoma; (2) primarily treated at the NKI. Dutch breast cancer radiology reports from Zhang et al.^21^ were also used in pretraining. This study was approved by the Institutional Review Board of the NKI with a waiver of informed consent (registration number: IRBd20-286).

During pretraining, all the rectal reports including radiology, pathology and endoscopy, were randomly divided into a 75:25 ratio on the patient level. Reports from 868 patients (75%) were used for pretraining, while the remaining 289 patients’ reports—entirely unseen during pretraining—were reserved for evaluating the pretrained models, see Figure 1. For the downstream survival prediction, patients were excluded if (1) their last follow-up status (mortality or alive) or tumor status (tumor present or absent) was unavailable; (2) radiology reports at diagnosis were missing, see Figure 1. Reports at diagnosis were defined as those recorded within 2 months of the diagnosis date. Following the exclusion process, a total of 399 radiology reports were utilized for training, 100 for internal validation, and 170 for testing in the overall survival analysis. For the disease-free survival task, 333 patients were designated for training, 84 for internal validation, and 125 for testing. The basic characteristics of the patients included in the survival tasks are presented in the Table 1 and Table 2.

**Table 1.** Summary of patient demographic and clinical characteristics for overall survival prediction.

|  |  | Train | Internal Val | Test | p-value |
| --- | --- | --- | --- | --- | --- |
| Age | < 65 | 231 (58%) | 62 (62%) | 105 (62%) | 0.29 |
|  | ≥ 65 | 168 (42%) | 38 (38%) | 65 (38%) |  |
| Gender | Female | 179 (45%) | 40 (40%) | 78 (46%) | 0.10 |
|  | Male | 220 (55%) | 60 (60%) | 92 (54%) |  |
| cT | 1 | 64 (16%) | 10 (10%) | 26 (15%) | 0.30 |
|  | 2 | 91 (23%) | 16 (16%) | 35 (21%) |  |
|  | 3 | 149 (37%) | 51 (51%) | 66 (39%) |  |
|  | 4 | 82 (21%) | 20 (20%) | 40 (24%) |  |
|  | missing | 13 (3%) | 3 (3%) | 3 (2%) |  |
| cN | 0 | 177 (44%) | 33 (33%) | 71 (42%) | 0.17 |
|  | N+ | 218 (55%) | 66 (66%) | 97 (57%) |  |
|  | missing | 4 (1%) | 1 (1%) | 2 (1%) |  |
| Survival Event | Alive | 291 (73%) | 72 (72%) | 114 (67%) | 0.36 |
|  | Mortality | 108 (27%) | 28 (28%) | 56 (33%) |  |
| Total |  | 399 | 100 | 170 |  |

The values in parentheses represent percentages. cT, baseline T staging. cN, baseline N staging. P-values were calculated using independent-sample t test or Mann-Whitney U test. Categorical variables were expressed as counts, and chi-squared test was used for frequency comparison.

**Table 2.** Summary of patient demographic and clinical characteristics for disease-free survival prediction.

|  |  | Train | Internal Val | Test | p-value |
| --- | --- | --- | --- | --- | --- |
| Age | < 65 | 205 (62%) | 45 (54%) | 78 (62%) | 0.15 |
|  | ≥ 65 | 128(38%) | 39 (46%) | 47 (38%) |  |
| Gender | Female | 149 (45%) | 36 (43%) | 55 (44%) | 0.37 |
|  | Male | 184 (55%) | 48 (57%) | 70 (56%) |  |
| cT | 1 | 51 (15%) | 11 (13%) | 19 (15%) | 0.86 |
|  | 2 | 62 (19%) | 20 (24%) | 23 (18%) |  |
|  | 3 | 139 (42%) | 33 (39%) | 45 (36%) |  |
|  | 4 | 73 (22%) | 19 (23%) | 35 (28%) |  |
|  | missing | 8 (2%) | 1(1%) | 3 (2%) |  |
| cN | 0 | 137 (41%) | 33 (39%) | 52 (42%) | 0.98 |
|  | N+ | 191 (57%) | 49 (58%) | 71 (57%) |  |
|  | missing | 5 (2%) | 2 (2%) | 2 (1%) |  |
| Survival Event | DFS | 214 (64%) | 49 (58%) | 78 (62%) | 0.60 |
|  | Non-DFS | 119 (36%) | 35 (42%) | 47 (38%) |  |
| Total |  | 333 | 84 | 125 |  |

The values in parentheses represent percentages. cT, baseline T staging. cN, baseline N staging. P-values were calculated using independent-sample t test or Mann-Whitney U test. Categorical variables were expressed as counts, and chi-squared test was used for frequency comparison.

**Figure 1.**
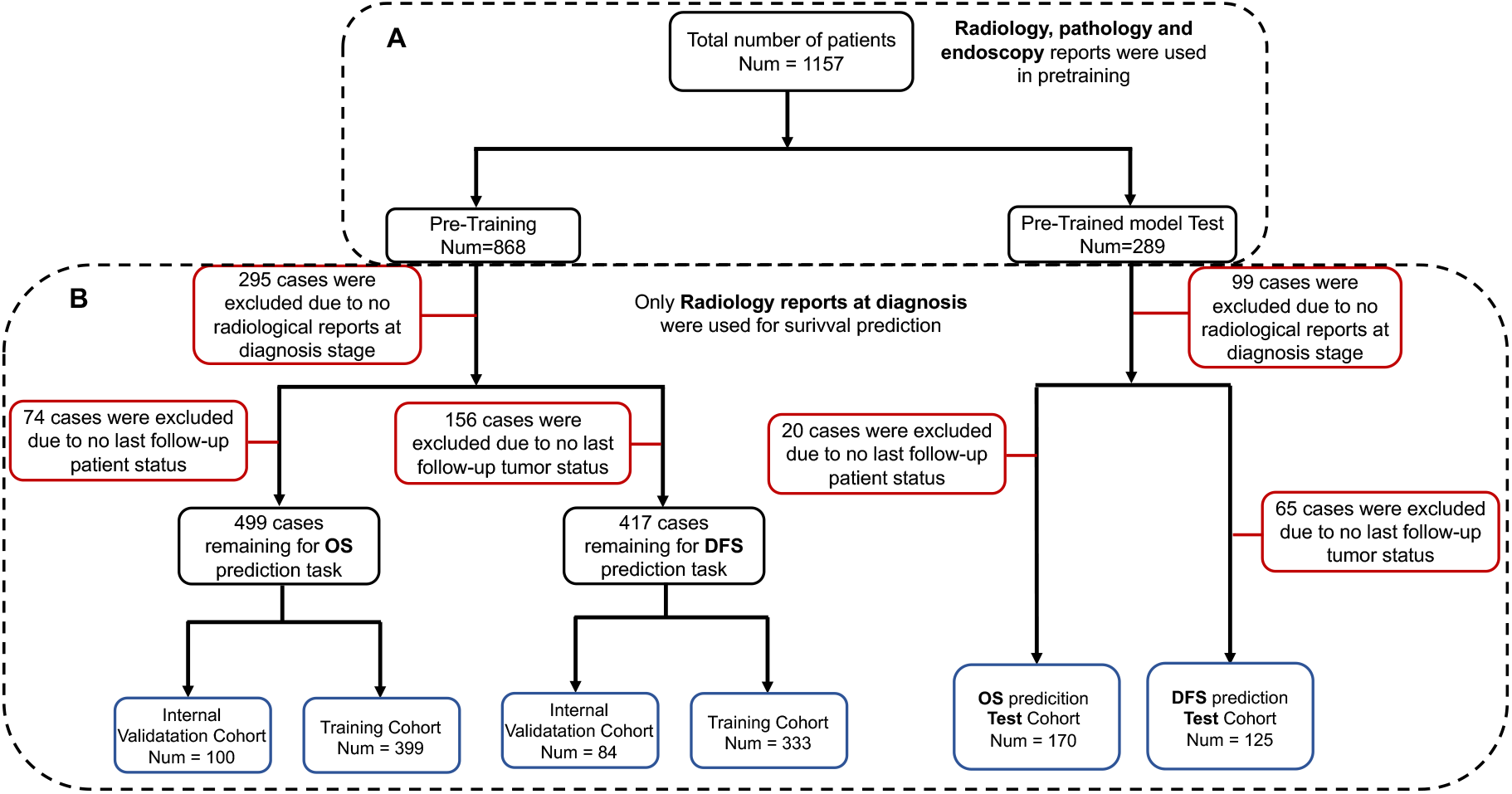
The flowchart of this study: A. The dataset used for pretraining. B. The exclusion process for OS and DFS tasks. Test dataset was not used in the pretraining.

### Pretrained NLP models

RoBERTa was trained using masked language modeling as shown in Figure 3. Our pretraining for RecRoBERT, BRecRoBERT and BRec2RoBERT involved randomly masking 15% of the input words, which is consistent with Zhang et al.^21^, where it was demonstrated that 15% masking probabilities in training yielded the highest accuracy in the test set. We evaluated the models’ comprehension of Dutch rectal cancer reports by comparing the prediction accuracy of different pretrained RoBERTa models using masked word prediction in the test set. The results are presented in Table 3.

**Table 3.** The accuracy of masked words prediction of 5 pretrained language models.

|  | RobBERT | MedRoBERTa.nl | RecRoBERT | BRecRoBERT | BRec2RoBERT |
| --- | --- | --- | --- | --- | --- |
| Accuracy | 0.34 | 0.50 | 0.82 | 0.81 | **0.85** |
| Top 5 Accuracy | 0.49 | 0.70 | 0.91 | 0.91 | **0.93** |
| Top 10 Accuracy | 0.54 | 0.76 | 0.93 | 0.93 | **0.94** |

Accuracy was determined by selecting the word with the highest probability. The Top 5 (Top 10) accuracy was computed by considering the top 5 (10) words for accuracy assessment.

RobBERT^32^, a RoBERTa-based language model exclusively trained on general Dutch texts, demonstrated the poorest masked words prediction performance on the test data, with an accuracy of 0.34. MedRoBERTa.nl^29^, pretrained on general clinical notes from Dutch hospitals, achieved an improved accuracy of 0.50, outperforming RobBERT. RecRoBERT, pretrained only on a limited number of rectal cancer reports, and BRecRoBERT, pretrained on the combination of breast and rectal cancer reports, showed similar accuracies of 0.80 and 0.81, respectively. BRec2RoBERT, which is essentially BRecRoBERT with a second round of pretraining with rectal cancer reports, achieved the highest accuracy of 0.85 among all the pretrained models in masked words prediction.

### Visualization of pretrained models

We present word clouds in Figure 2 for all reports as well as for radiology-specific reports. The word clouds show different word frequencies in radiology reports; for example, terms like ‘MRI’, ‘CT’ (Computed Tomography), and ‘radiologist’ appeared more frequently in radiology-specific reports. We visualized the pretrained models using one short example report to gain more insight into what the model learned during the pretraining process. Heatmaps in Figure 4 were generated using the attention weights from 12th layer of the first head of RoBERTa to show the correlations between words from the given MRI report. RobBERT and MedRoBERTa.nl exhibited limitations during tokenization, as they were unable to encode the term like ‘semicircular’ as a single token. This shortcoming compromises their capacity to accurately represent key terminology used in the morphological description of rectal tumors. For RecRoBERT, some word pair correlations lacked clinical relevance, for instance, ‘semicircular’ has strongest association with ‘/’ and ‘-’. BRecRobBERT and BRec2RobBERT showed more clinically meaningful word correlations. An interactive visualization^33^ in Figure 5 generated by BRecRoBERT showed that ‘Morphology’ is most closely associated with ‘semicircular’, indicating an improved understanding of rectal cancer reports.

**Figure 2.**
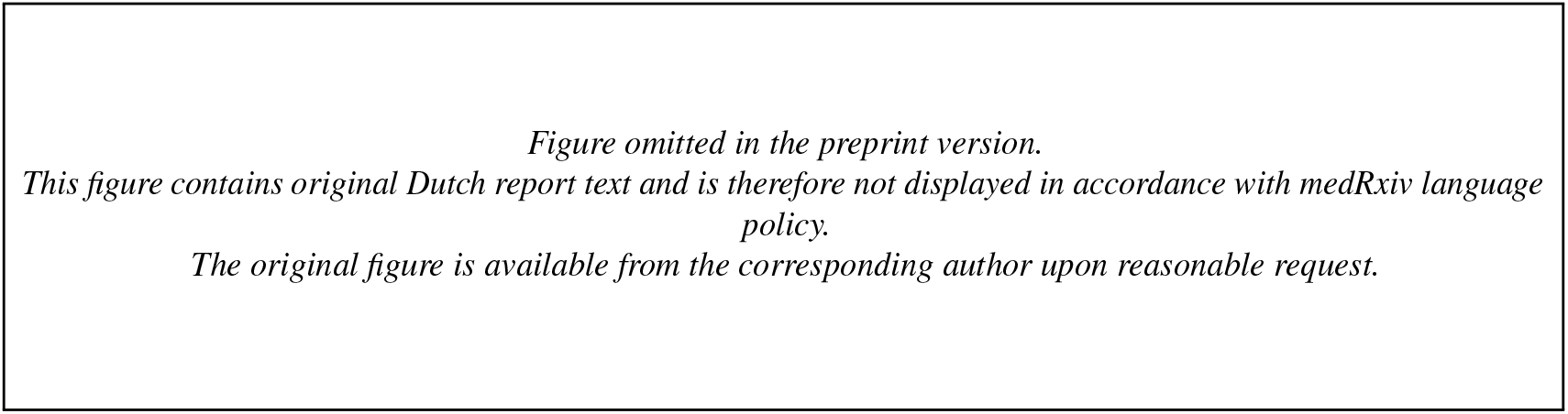
Word clouds. The image at the left describes the word frequencies of all three types reports (Radiology, Pathology, and Endoscopy), while the right image focuses exclusively on radiology reports.

**Figure 3.**
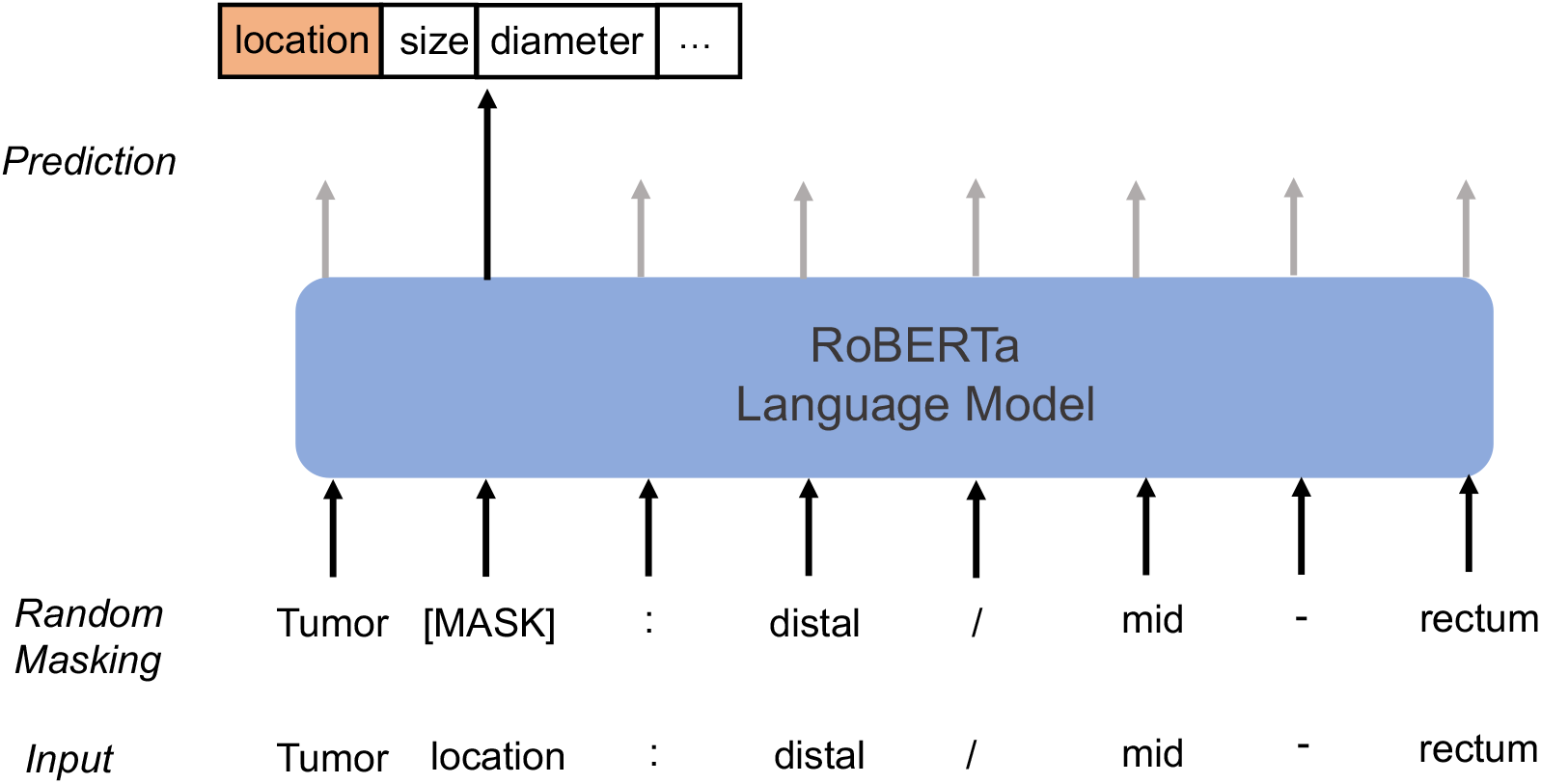
Masked words prediction with RoBERTa.

**Figure 4.**
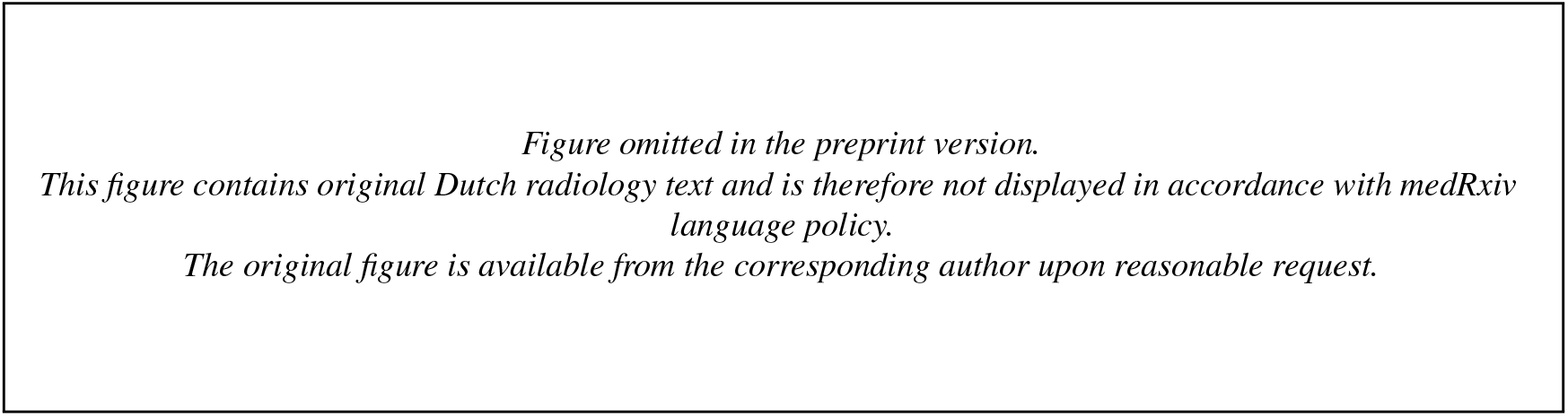
Heatmaps generated from the attention weights of five different pretrained RoBERTa models, based on a specific MRI report.

**Figure 5.**
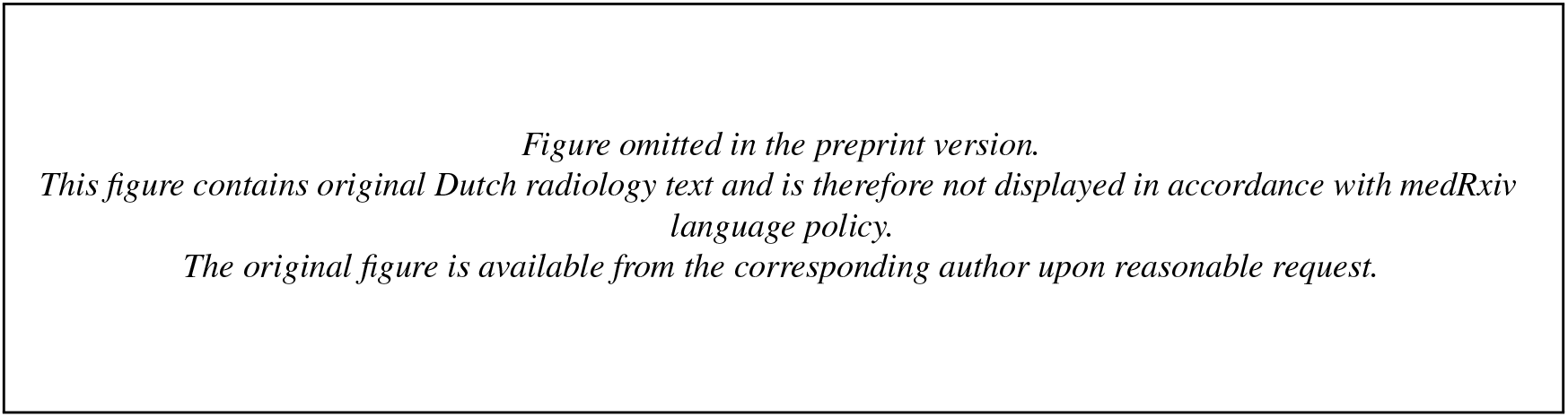
Interactive visualization of words correlation using BRecRoBERT

### Survival Prediction

For downstream survival prediction, we focused on two endpoints: overall survival (OS) and disease-free survival (DFS). Five deep learning (DL) models were trained on features extracted from five pretrained RoBERTa models to generate risk scores. The risk groups (high vs. low) were then stratified using the threshold defined by the median risk score obtained from the internal validation set. The resulting concordance index (C-index) values are shown in Table 4, with corresponding Kaplan-Meier (KM) curves in Figure 6 and 7 along with hazard ratios and Log-rank P values to assess the models’ performance.

**Table 4.** The C-index and HR for five pretrained RoBERTa models in OS and DFS tasks.

|  | RobBERT | MedRoBERTa.nl | RecRoBERT | BRecRoBERT | BRec2RoBERT |
| --- | --- | --- | --- | --- | --- |
| OS C-index (95% CI) | 0.59 (0.50, 0.68) | 0.56 (0.47, 0.65) | 0.59 (0.50, 0.68) | **0.65 (0.57, 0.73)** | 0.63 (0.55, 0.71) |
| DFS C-index (95% CI) | 0.65 (0.56, 0.74) | 0.59 (0.50, 0.66) | 0.61 (0.51, 0.70) | **0.71 (0.64, 0.78)** | 0.69 (0.60, 0.77) |
| OS HR (95% CI) | 1.66 (0.98-2.81) | 1.33 (0.75-2.34) | 1.64 (0.92-2.93) | **2.67 (1.51-4.72)** | 2.34 (1.32-4.16) |
| Log-rank P (OS) | 0.06 | 0.32 | 0.09 | <0.001 | 0.003 |
| DFS HR (95% CI) | 1.88 (1.05-3.37) | 1.87 (1.02-3.42) | 1.49 (0.83-2.66) | **3.81 (2.05-7.06)** | 2.91 (1.62-5.25) |
| Log-rank P (DFS) | 0.03 | 0.04 | 0.18 | <0.001 | <0.001 |

95% CI: 95% confidence intervals. OS: overall survival. DFS: disease-free survival. Log-rank p: Log-rank p value. HR: Hazard Ratio.

**Table 5.** Pretrained models and their coropas.

| Pretrained models | Pretraining Text | Source |
| --- | --- | --- |
| RobBERT | Dutch section of the OSCAR corups (39 GB) | Delobelle et al.<sup>32</sup> |
| MedRoBERTa.nl | Text data from Dutch hospital notes (12.3 GB) | Verkijk et al.<sup>29</sup> |
| RecRoBERT | Reports from 868 rectal cancer patients (24.1 MB) | We developed this model |
| BRecRoBERT | Reports from 868 rectal cancer patients and 37,517 breast cancer patients (24.1 MB + 86.61 MB) | We developed this model |
| BRec2RoBERT | Retrain BRecRoBERT using reports from 868 rectal cancer patients (24.1 MB) | We developed this model |

**Figure 6.**
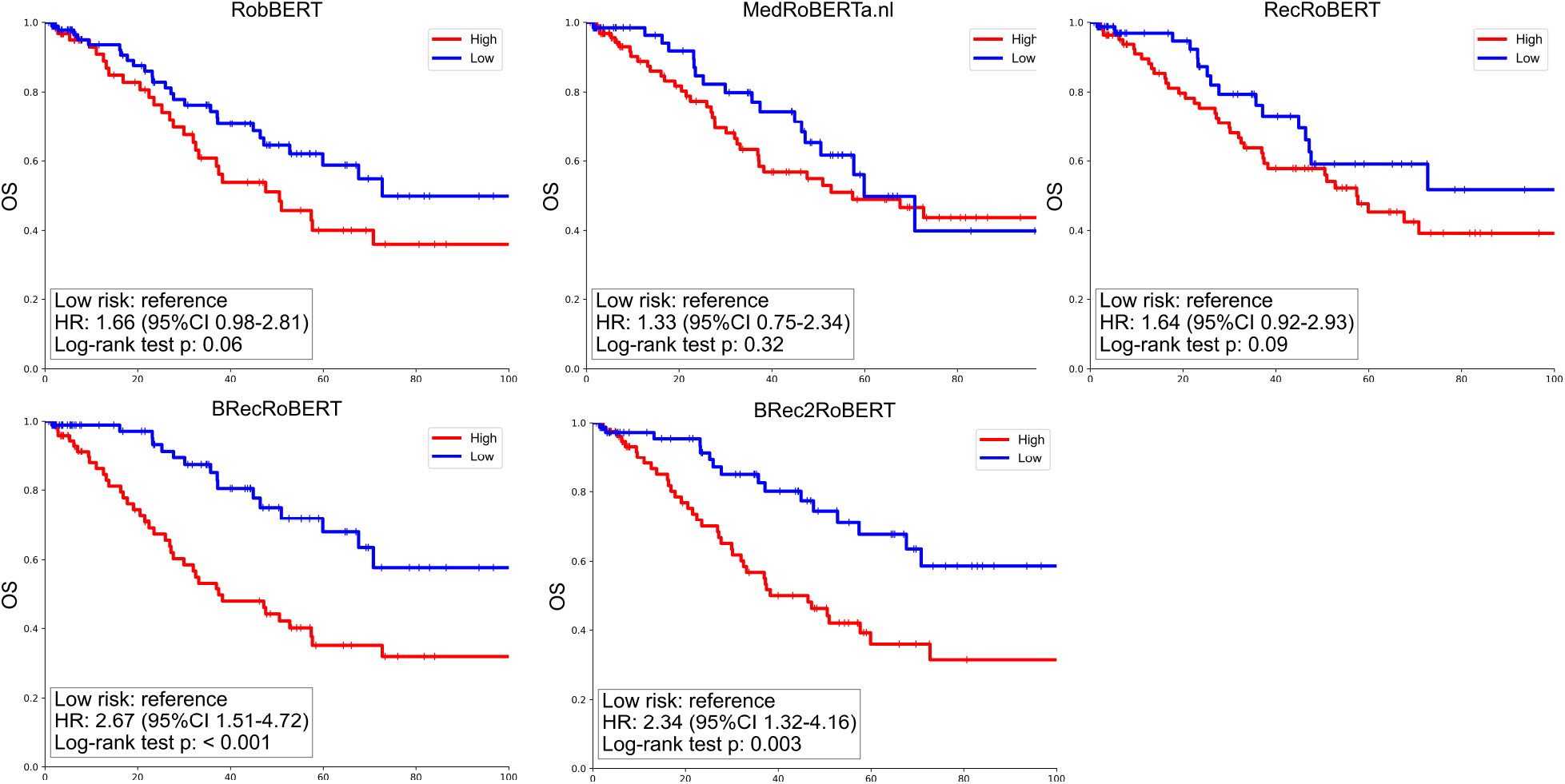
Kaplan-Meier analysis for OS by deep learning-based risk score in the test set.

**Figure 7.**
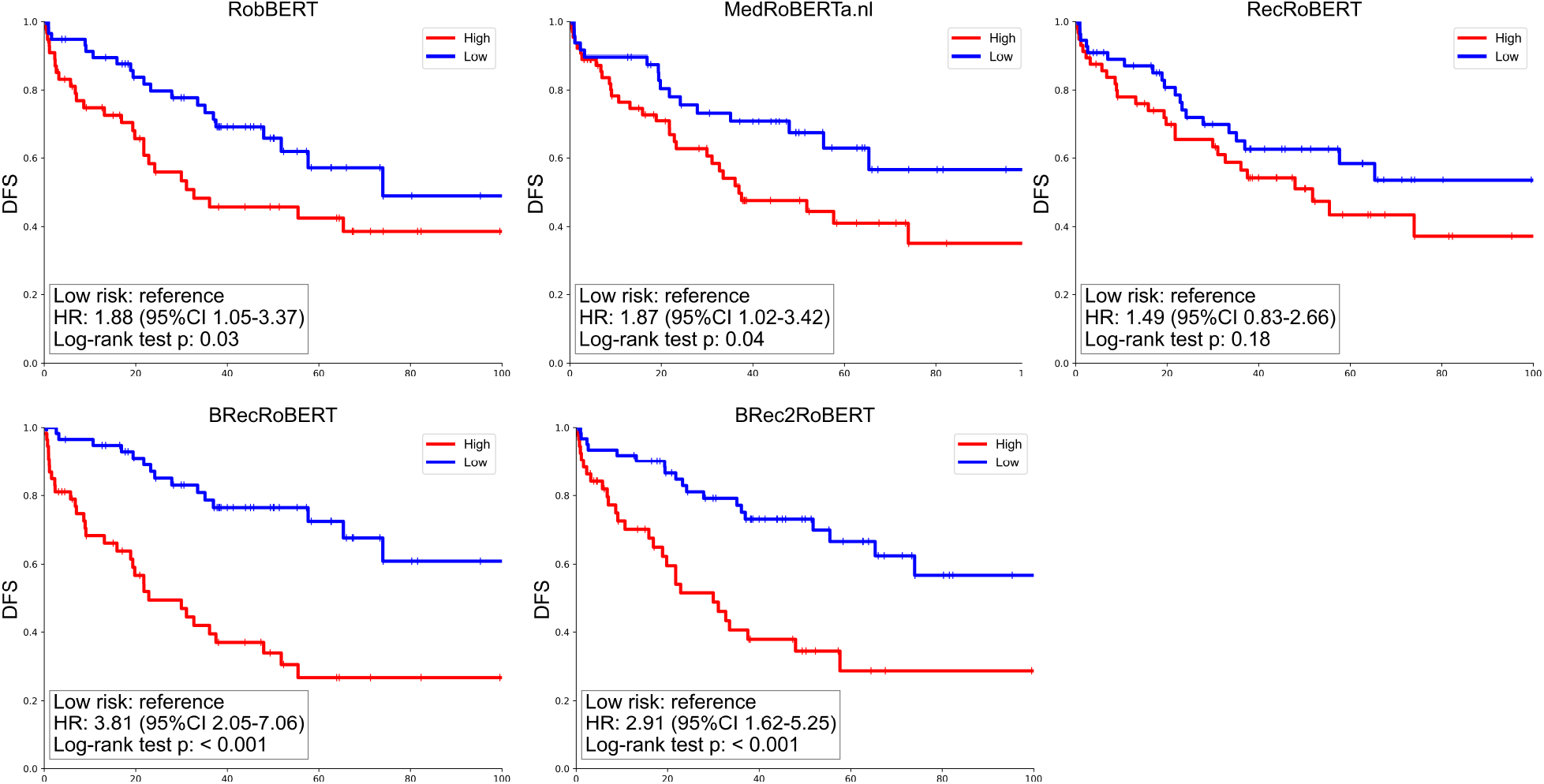
Kaplan-Meier analysis for DFS by deep learning-based risk score in the test set.

In both tasks, BRecRoBERT achieved the highest C-index, with values of 0.65 (0.57-0.73) for OS and 0.71 (0.64-0.78) for DFS, respectively. Furthermore, it achieved hazard ratios (HR) of 2.67 (1.51-4.72) for OS and 3.81 (2.05-7.06) for DFS. Both log-rank test p-values were below the 0.05 threshold. RobBERT and MedRoBERTa.nl performed poorly in both tasks. Although RecRoBERT demonstrated relatively high accuracy in masked word prediction, this advantage did not carry over to both the survival prediction applications. While BRec2RoBERT excelled in masked word prediction, it did not achieve the best results in survival prediction but still outperformed RobBERT, MedRoBERTa.nl and RecRoBERT.

To better understand the differences between the low and high risk groups, a text analysis was conducted using scatter text plots^34^ on the test set for both both OS and DFS tasks (BRecRoBERT), see Figure 10 and 11. In OS analysis, ‘ct1’ (clinical T staging 1) and ‘n0’ (N staging 0) were strongly linked to the low-risk group, while ‘liver metastases’ was associated with high-risk patients. For DFS, other than ‘liver metastases’, the word ‘both-sided’ was also highly associated with high risk.

### N-year Survival

The 1-year, 3-year, and 5-year survival prediction performance, based on patient risk scores from the deep learning model, was further assessed using the area under the receiver operating characteristic curve (AUC), as illustrated in Figures 8 and 9. BRecRoBERT-derived embeddings exhibited better performance in predicting 1-year overall survival (OS) with an AUC of 0.73 (0.60, 0.85) and 3-year OS with an AUC of 0.67 (0.56, 0.77), while achieving second place for 5-year OS predictions, with an AUC of 0.66 (0.54, 0.78). Furthermore, BRecRoBERT consistently outperformed other models in predicting DFS across all evaluated N-year intervals.

**Figure 8.**
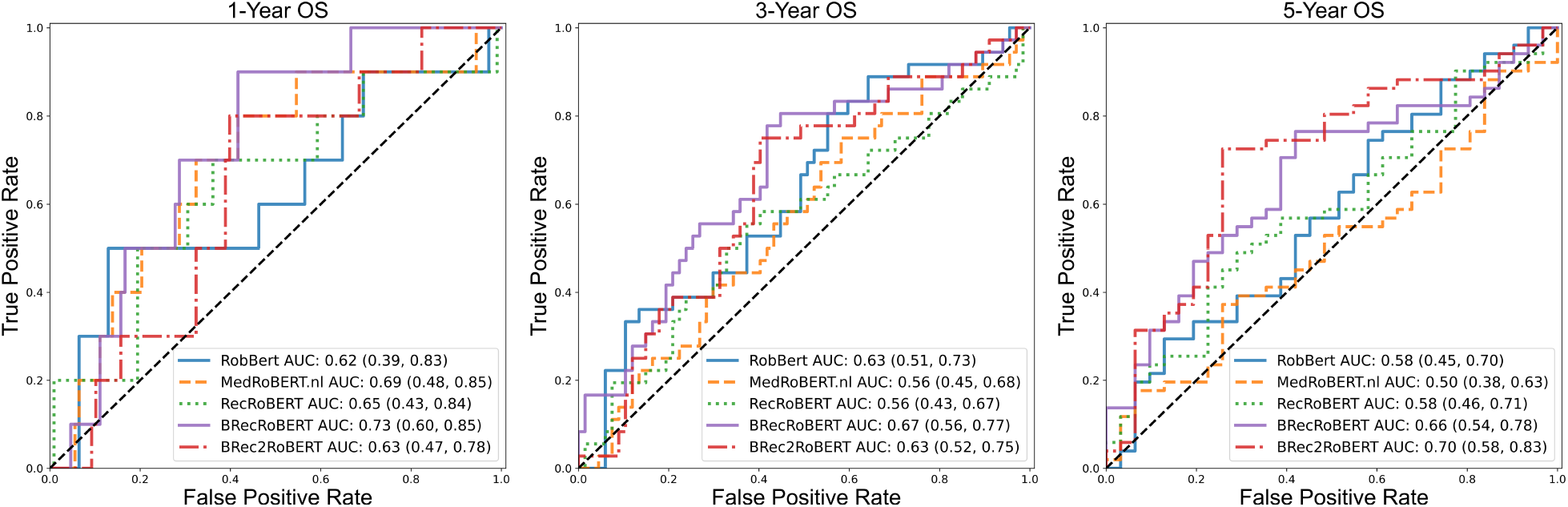
Receiver operating characteristic (ROC) curves for the predicted risk on N-year OS prediction.

**Figure 9.**
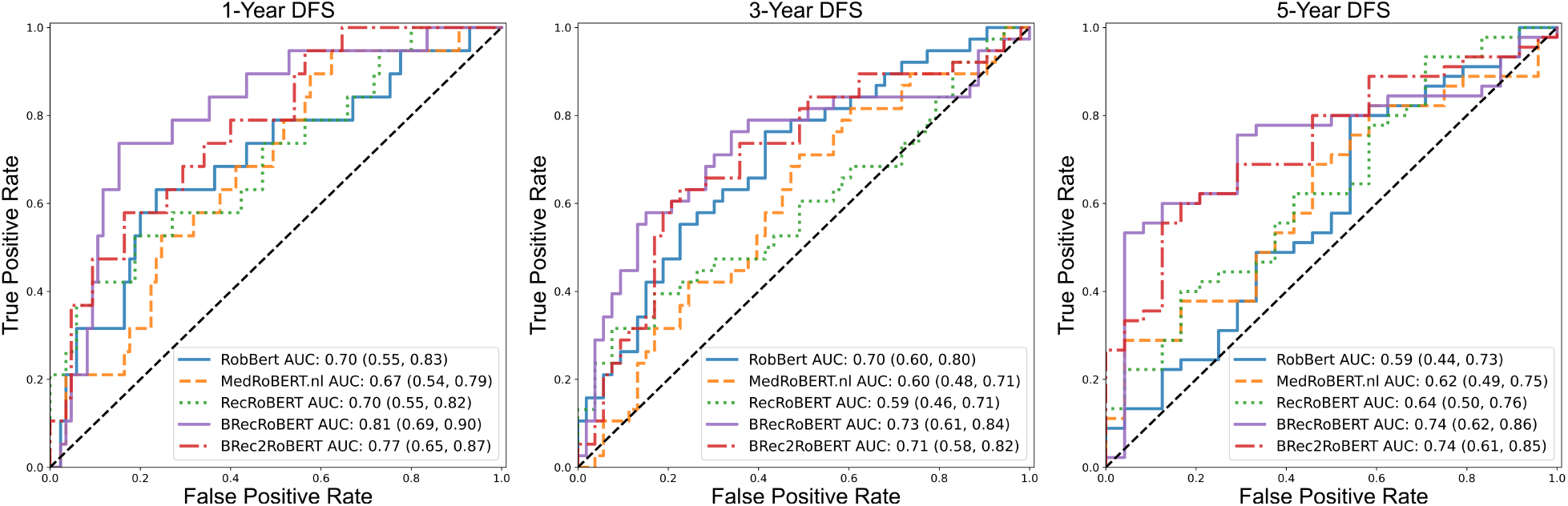
Receiver operating characteristic (ROC) curves for the predicted risk on N-year DFS prediction.

**Figure 10.**
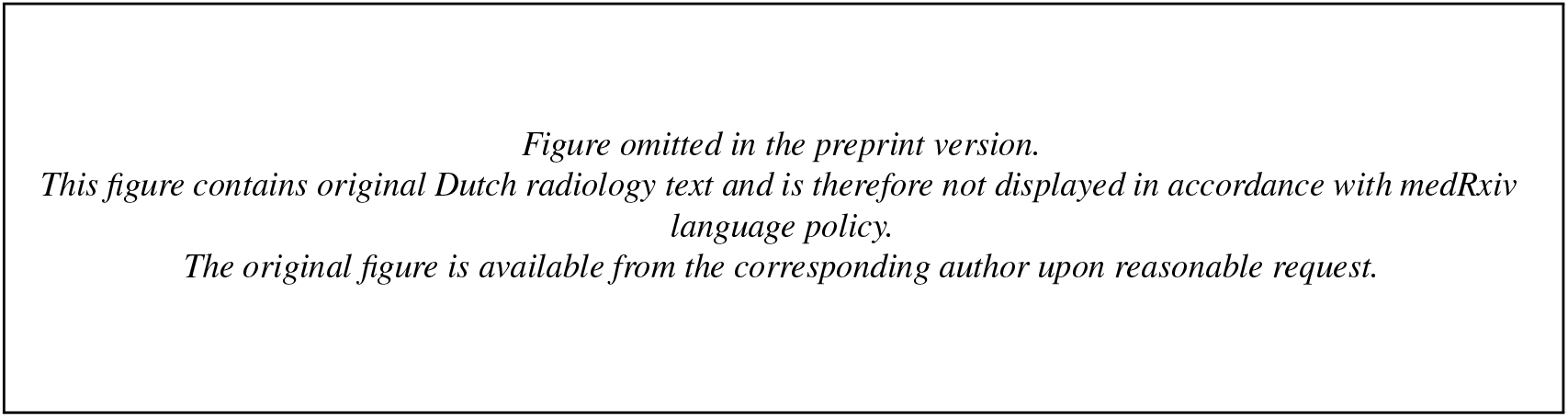
Scatter text analysis for OS in the test set.

**Figure 11.**
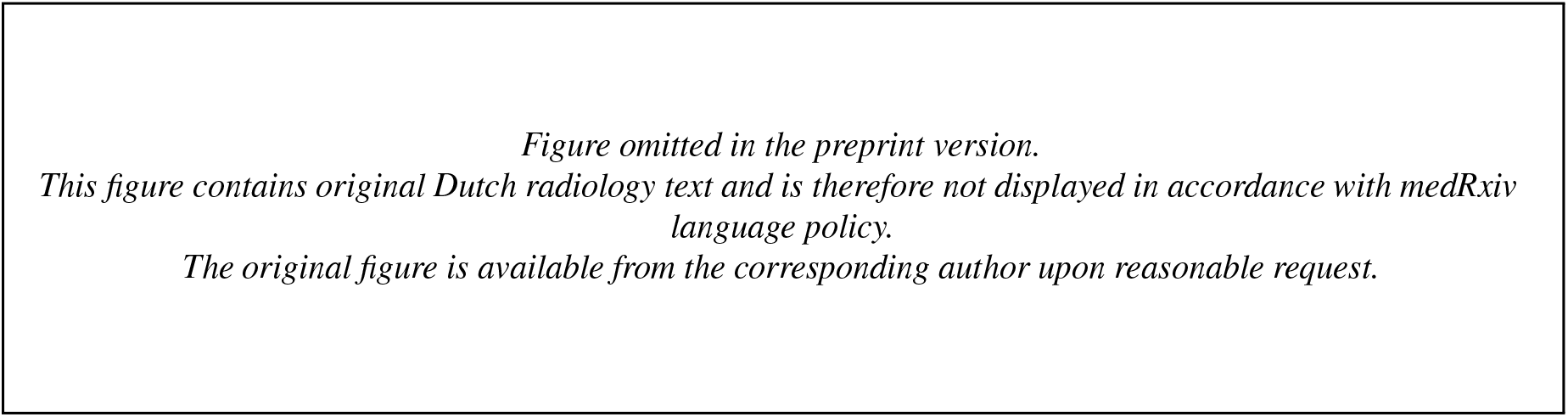
Scatter text analysis for DFS in the test set.

**Figure 12.**
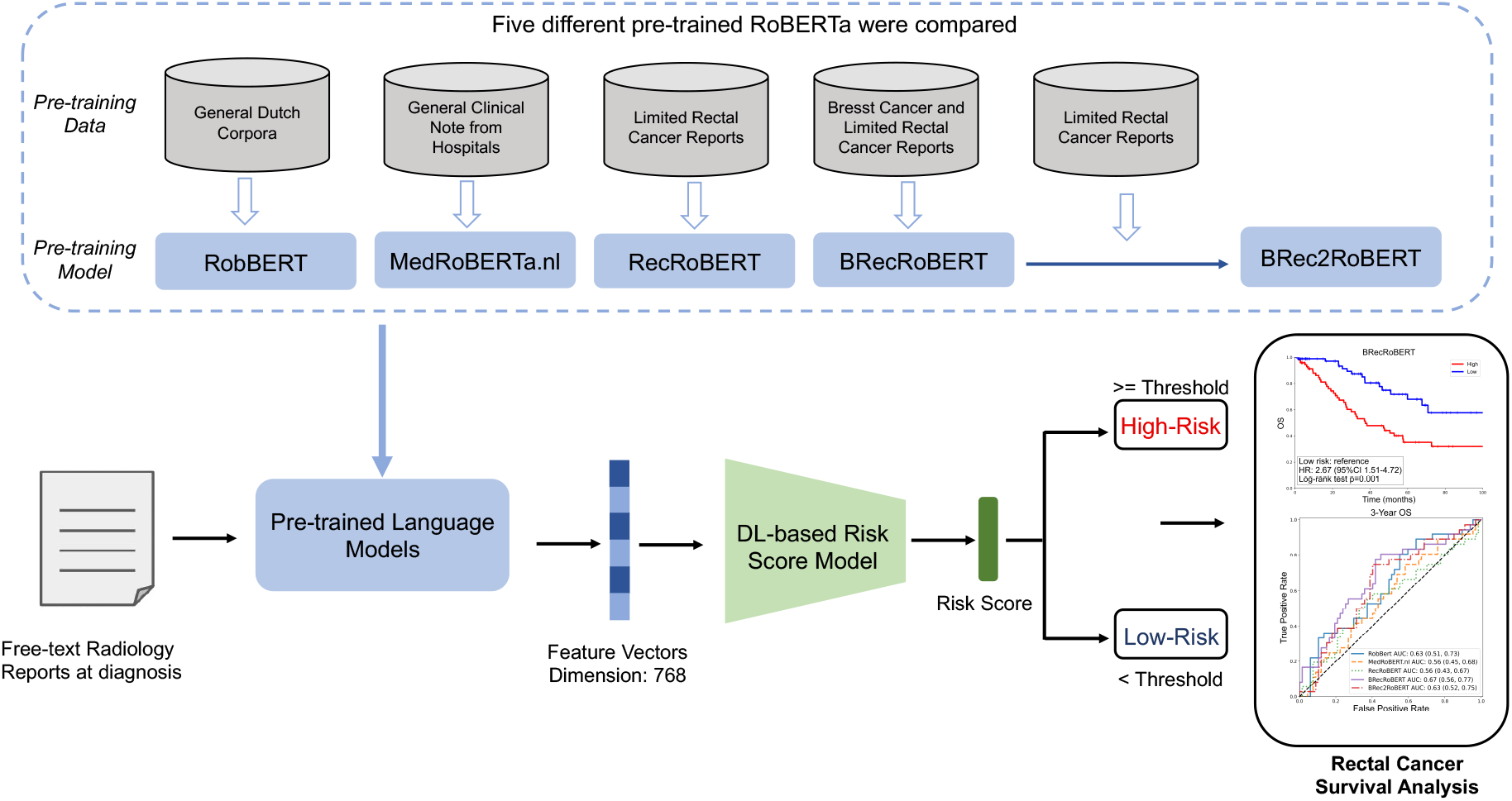
Graphical Abstract

## Discussion

In this study, we developed BRecRoBERT, a RoBERTa model pretrained on a combination of Dutch breast and rectal cancer reports. BRecRoBERT significantly improved the prediction of survival outcomes for rectal cancer patients, outperforming models (RobBERT and MEdRoBERTa.nl) pretrained on general Dutch corpora. We emphasized the importance of using pretrained language models with domain-specific knowledge for the downstream tasks. Moreover, we found that applying data from related domains can improve the quality of feature vectors, especially in cases where domain-specific data is scarce. RecRoBERT, trained exclusively with rectal cancer text, showed limited performance in survival prediction, even though it was successful in masked words prediction. Integrating breast cancer radiology reports during pretraining, BRecRoBERT achieved better survival prediction results compared to RecRoBERT.

Kim et al.^20^ utilized ClinicalBERT to extract features from MRI reports of rectal cancer patients for survival prediction. However, ClinicalBERT lacks understanding in rectal cancer reports. Frequently used terms like ‘rectum’, ‘distal’ and ‘morphology’ were out-of-vocabulary words and they were tokenized using subword tokenization based on the WordPiece algorithm, see in Supplementary. Tokenization is a crucial process for language models, as it transforms words into numerical representations by associating words with token IDs. Out-of-vocabulary words or phrases were broken into subwords, which can lead to gaps in understanding and a failure to recognize relevant concepts in the data^35^. Also, no clinically significant word correlations were detected when using ClinicalBERT, as shown in Figure S1. Similarly, RobBERT and MedRoBERTa.nl without rectal cancer knowledge showed poor performance in both masked word prediction and survival tasks, which further demonstrated that domain-specific models tend to outperform more general models on tasks within their respective domains. Verkijk et al.^29^ have pointed out that with sufficient in-domain data, training a model from scratch appears to be more effective than extending the pretraining of an existing model. RecRoBERT, which was pretrained from scratch on a limited set of Dutch cancer records, performed well in masked word prediction but failed to generate high-quality feature embeddings for survival prediction. This highlighted that strong performance in masked word prediction does not always guarantee success in more complex downstream applications and the importance of sufficient data during the pretraining. Dutch breast and rectal cancer reports, though focused on different types of cancer, may contain overlapping clinical concepts, such as tumor staging, treatment strategies, and diagnostic procedures. For instance, we observed that the word ‘rectum’ exists in the vocabulary of pretrained model using only breast cancer records by Zhang et al.^21^. Incorporating a substantial number of Dutch breast cancer reports during pretraining improved survival prediction tasks for rectal cancer, indicating that pretrained models can draw on knowledge from a related domain when truly in-domain data are scarce. The pretrained RoBERTa model may benefit from data on other cancer types, such as colon, prostate, and bladder cancer, due to the inherent similarities in their associated reports. BRec2RoBERT, with the highest masked word prediction accuracy, did not outperform BRecRoBERT in survival prediction. The reason for this might be that the fine-tuning process with limited rectal cancer reports can increase the inherent risk of over-specialisation in training data.

In downstream survival prediction, disease-free survival appears to be an easier task compared to overall survival, as evidenced from the improved C-index and hazard ratios. This aligns with the study by Wei et al.^36^. Overall survival remains more challenging to predict because while patients may remain disease-free, they can still die from other causes over the long term, and factors affecting mortality may not be documented in radiology reports at the time of diagnosis. In the scatter text analysis for both OS and DFS, the deep learning-based risk score model identified a strong correlation between ‘liver metastasis’ and ‘metastasis’ with high-risk patients, suggesting worse survival outcomes. These findings are in line with established clinical evidence^37^. The model associated keywords like ‘cT1’ (clinical stage 1) and ‘N0’ (N staging 0) with low risk in the OS task, suggesting better overall survival outcomes. This is consistent with the findings of Bogveradze et al.^38^, where cT1-2 and N0 were highlighted as key indicators of low-risk patients. Certain terms, such as ‘anorectal’, ‘anorectal junction’, and ‘above’, appeared to be correlated with low-risk patients in both tasks, despite not being directly indicative of clinical risk levels. This could be due to the contextual usage of these terms in reports associated with early-stage or less severe cases. These terms may frequently occur in patients with favorable prognoses, and the model therefore learns to associate them with low risk.

Beyond unstructured reports, recent studies have demonstrated that integrating additional modalities such as structured texts, imaging biomarkers, and laboratory test results can improve predictive performance, particularly in tasks like liver metastase and anastomotic leakage prediction^39,40^. Multimodal data provide complementary information: structured data offer patient demographics and medical history, imaging biomarkers capture spatial and morphological features, while laboratory data reflect physiological and biochemical states. Together, they enable a more comprehensive understanding of the patient’s condition. Incorporating such multimodal inputs into deep learning models holds great potential to further support clinical decision-making in rectal cancer care.

## Data Availability

All data used in this study are not publicly available due to privacy and ethical restrictions. Access may be granted upon reasonable request and with appropriate institutional approval.

https://github.com/Liiiii2101/Dutch-RoBERTa-for-Rectal-Cancer-Reports

## Limitations of the study

This study has several limitations. First, all the data was retrospectively collected and conducted at a single Dutch medical institute, which may limit the generalizability of our findings. Clinical documentation practices, linguistic expressions, and terminology can vary significantly across different medical centers. These differences may impact model performance when applied in broader or multilingual settings. Our future work will focus on external validation using data from multiple centers and exploring multilingual pretraining strategies to improve the robustness and applicability of the model. Second, a very limited amount of data was available in the survival prediction tasks. The study could therefore benefit from a larger cohort. Third, only breast cancer reports were combined and investigated for pretraining models due to the data availability. However, other cancer reports such as colon cancer, prostate cancer could potentially help improve the quality of the pretrained models given that they are closer in terms of anatomical structures.

## STAR ⋆ Methods

Detailed methods are provided in the online version of this paper and include the following:

- KEY RESOURCES TABLE
- RESOURCE AVAILABILITY

- Lead contact
- Materials availability
- Data and code availability

- EXPERIMENTAL MODEL AND SUBJECT DETAILS

- Data collection

- METHOD DETAILS

- Pretrained Language Models
- Survival Prediction
- Training Details

- QUANTIFICATION AND STATISTICAL ANALYSIS

## SUPPLEMENTAL INFORMATION

Supplemental information can be found in the supplementary material.

## ACKNOWLEDGMENTS

This project has received funding from the European Union’s Horizon 2020 Research and Innovation Programme under the Marie Skłodowska-Curie grant agreement No 857894. The study was supported by the Research High Performance Computing (RHPC) facility of the Netherlands Cancer Institute.

## AUTHOR CONTRIBUTIONS

L C drafted the manuscript. L C and R B acquired the data. L C, T Z, and J B contributed to the data analysis and interpretation. T Z, J B, and L C contributed to the experimental design and manuscript revision. J B, J T, and R B supervised the project.

## STAR ⋆ Methods

### KEY RESOURCES TABLE

| REAGENT or RESOURCE | SOURCE | IDENTIFIER |
| --- | --- | --- |
| Source code | This paper | https://github.com/Liiiii2101/Dutch-RoBERTa-for-Rectal-Cancer-Reports |
| Supplementary Figures and Tables | This paper | Supplementary Files |
| Software and Algorithms |  |  |
| Python 3.9.12 | Python | https://www.python.org/downloads/release/python-3912/ |
| torch 2.1.2 | PyTorch | https://pytorch.org/ |
| huggingface-hub 0.20.3 | Hugging Face | https://huggingface.co/ |
| transformers 4.37.0 | Hugging Face | https://huggingface.co/docs/transformers/index |
| tokenizers 0.15.1 | Hugging Face | https://huggingface.co/docs/tokenizers/index |
| scipy 1.13.0 | Scipy | https://scipy.org/ |
| numpy 1.24.4 | NumPy | https://numpy.org/ |
| bertviz 1.4.0 | BertViz | https://pypi.org/project/bertviz/ |
| scattertext 0.1.19 | ScatterText | https://github.com/JasonKessler/scattertext |
| lifelines 0.28.0 | lifelines | https://pypi.org/project/lifelines/ |
| scikit-learn 1.4.2 | scikit-learn | https://scikit-learn.org/1.5/whats_new/v1.4.html |

### RESOURCE AVAILABILITY

#### Lead contact

Further information and requests for resources should be directed to and will be fulfilled by the lead contact, Jonas Teuwen

#### Materials availability

This study did not generate new unique reagents

#### Data and code availability

The original data is private and is not publicly available to guarantee protection of patients’ privacy. Code can be accessed at https://github.com/Liiiii2101/Dutch-RoBERTa-for-Rectal-Cancer-Reports

### EXPERIMENTAL MODEL AND SUBJECT DETAILS

#### Data Collection

In total, 57 different radiologists authored the radiology reports. The diagnostic radiology reports were defined as those that include MRI and/or CT, and/or Positron Emission Tomography (PET) findings generated within two months within the diagnosis date.

### METHOD DETAILS

#### Pretrained Language Models

In this study, we compared five different pretrained langauge models including RobBERT, MedRoBERTa.nl, RecRoBERT, BRecRoBERT and BRec2RoBERT in both masked words prediction tasks and their generated feature vectors for survival prediction. RobBERT and MedRoBERTa.nl are publicly available. RecRoBERT, BRecRoBERT and BRec2RoBERT were implemented based on RoBERTa. The pretraining procedure consisted of a few steps. First, the words in the reports were tokenized and mapped to corresponding tokens. Roberta uses a Byte Pair Encoding (BPE) tokenizer, where the text is tokenized to variable-length byte n-grams^41^. The benefit of the byte-level tokenizer lies in its ability to encode any input text, even if it is out-of-vocabulary. RobBERT and MedRoBERTa.nl have vocabulary sizes of 40,000 and 52,000, respectively, while we pre-defined the vocabulary size of our in-house model to 30,522, consistent with the approach used in Zhang et al.^21^. These tokens were then passed through an embedding layer, and the resulting embeddings were used as inputs to the transformer encoder layers. The tokens were processed iteratively across multiple encoder layers. Following this, a decoder layer predicted the attributes of each input token. 12 layers (L=12) of transformer blocks were employed, each with a hidden size of 768 (H=768) and 12 parallel self-attention heads (h=12). The self-attention mechanism was divided into three components: query (Q), key (K), and value (V). The Q and K were combined using the scaled dot-product method to compute attention weights, which were then normalized using the softmax function. Lastly, the weighted sum of these weights and V was computed to obtain refined feature representations for each token. Masked language modeling was employed during pretraining to enhance the model’s ability to understand Dutch reports. Different pretrained models were using different dataset. RecRoBERT was trained solely on the rectal cancer dataset, while BRecRoBERT was trained on both breast and rectal cancer datasets. BRec2RoBERT underwent an initial training phase on both breast and rectal cancer datasets, followed by additional training on the rectal cancer dataset alone. Pretraining is an unsupervised learning process in which words in the input sequences were randomly masked with probabilities of 15%, in line with Zhang et al.^21^. The model was then trained to reconstruct the masked words.

#### Embedding Extraction

At the beginning of each input sequence, a special classification token [CLS] is added. After processing through the full transformer stack, the final hidden state corresponding to the [CLS] token is used as a summary representation of the entire input sequence. In this work, we extract this [CLS] embedding from the final layer of a frozen pre-trained RoBERTa model and use it as a global feature vector representing the radiology report.

#### Survival Prediction

For survival prediction tasks, we employed a multiplelayer perceptron (MLP), inspired by Jiang et al. and DeepSurv model^6,42^, for risk score prediction. The model utilizes feature vectors extracted from pretrained language models as input. These vectors were passed through an encoder, implemented as a fully connected layer, which transforms the input into 256-dimensional features. Batch normalization and dropout regularization are applied to prevent overfitting. The final fully connected layer reduced the feature space, producing the risk score as the model’s output. As the loss function, we used the Cox partial likelihood (which has been shown to be effective in survival prediction studies^6,42,43^), defined as:

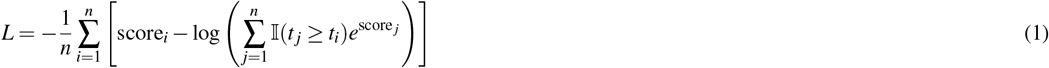

where: *score*_*i*_ is the predicted risk score for individual i, *t*_*i*_ is the observed time for individual i, 𝕀 (*t* _*j*_ ≥ *t*_*i*_) is an indicator function that is 1 if *t* _*j*_ ≥ *t*_*i*_ and 0 otherwise, and n is the number of individuals.

#### Training Details

During the unsupervised pretraining of the language models (RoBERTa), training was conducted for 10 epochs with a batch size of 32 and a learning rate of 1e-4, using the AdamW optimizer to update the parameters. For training the risk score prediction model, a batch size of 4 and an initial learning rate of 1e-5 were employed. Adam was used as the optimizer. To minimize the risk of overfitting, L1 and L2 regularization were utilized and the early stopping patience is 10. We trained the model for 100 epochs. The optimal model checkpoints were determined by selecting the epoch that yielded the highest C-index values for overall survival (OS) or disease-free survival (DFS) tasks on the internal validation set, with the OS and DFS models trained and evaluated separately. The threshold for categorizing high and low-risk groups was determined using the median risk scores from the internal validation set. To ensure reproducibility of the risk score deep learning model, we use a fixed random seed (seed=42). For a fair comparison of different embeddings, identical experimental setups, including data splits and evaluation metrics, are maintained. All models were trained on a single NVIDIA RTX A6000 graphics processing unit (GPU), with 48 Gigabytes of GPU memory.

### QUANTIFICATION AND STATISTICAL ANALYSIS

#### Statistical Analysis

Statistical analysis was performed using Python 3.9. Accuracy was used to evaluate the performance of the masked words prediction. Concordance index (C-index) were used to evaluate the predictive accuracy of survival models and C-index takes the censored data into account. Area under the receiver operating characteristic curve (AUC) were used to measure N-year survival. 95% confidence intervals were computed using the bootstrap method with 1,000 replications. Kaplan-Meier analysis and the log-rank test were employed to compare survival differences between the groups. Additionally, a Cox proportional hazards model was applied based on this grouping. The Kaplan-Meier method, the log-rank test and Cox proportional hazards model were done using the lifelines package version 0.28.0.

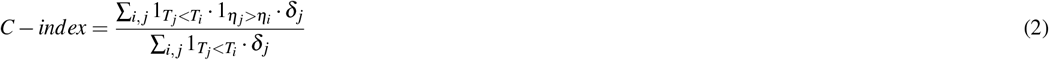

with *T*_*i*_ is the actual survival times for individual i. *η*_*i*_ the risk score of individual *i*, 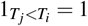 if *T*_*j*_ *< T*_*i*_, else 0. 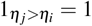 if *η*_*j*_ *< η*_*i*_, else 0. *δ* _*j*_ is the an indicator for whether the pair i, j is valid for comparison.

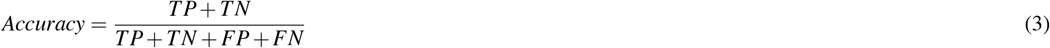

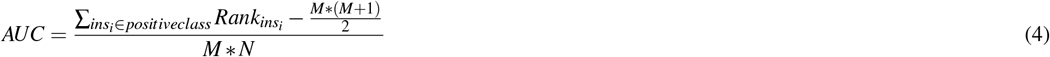

For AUC, M, N are the number of positive samples and negative samples. 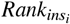 is the serial number of sample i. 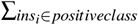 is adding up he serial numbers of the positive cases.

## Highlights

- Pretraining language models with domain-specific knowledge improves their performance in clinical applications.
- Using data from related domains can refine features for clinical tasks, particularly when domain-specific data is scarce.
- BRecRoBERT, pretrained on rectal and breast cancer texts, demonstrated the best features for predicting OS and DFS.

