## Supplementary for "Predicting Rectal Cancer Patient Survival with Dutch Radiology Reports using Natural Language Processing (NLP): The Role of Pretrained Language Models"

### Tokenization

The example below illustrates tokenization results for one MRI report using five pre-trained Dutch models. The original Dutch report excerpt is omitted in compliance with medRxiv policy. The report was also translated into English and tokenized using ClinicalBERT for comparison.

- **Original Dutch report:** *Omitted in compliance with medRxiv policy.*
- **English translation:** “Tumor location: distal/mid-rectum, starting approximately 3 cm above the anorectal junction, length approximately 3.8 cm – Morphology: solid, semicircular from approximately 8–12 o’clock.”
- **RobBERT tokens (count = 57):** [<s>, TOKEN_1, TOKEN_2, TOKEN_3, ..., </s>]
- **MedRoBERTa.nl tokens (count = 44):** [<s>, TOKEN_1, TOKEN_2, TOKEN_3, ..., </s>]
- **RecRoBERT tokens (count = 43):** [<s>, TOKEN_1, TOKEN_2, TOKEN_3, ..., </s>]
- **BRecRoBERT tokens (count = 43):** [<s>, TOKEN_1, TOKEN_2, TOKEN_3, ..., </s>]
- **BRec2RoBERT tokens (count = 43):** [<s>, TOKEN_1, TOKEN_2, TOKEN_3, ..., </s>]
- **ClinicalBERT tokens (count = 47):** [[CLS], tumor, location, :, dis, ##tal, /, mid, -, re, ##ctum, ,, starting, approximately, 3, cm, above, the, ano, ##rec, ##tal, junction, ,, length, approximately, 3, ., 8, cm, -, mor, ##phology, :, solid, ,, semi, ##cir, ##cular, from, approximately, 8, -, 12, o, ', clock, [SEP]]

**Supplementary Table 1.** The C-index and HR for Bag-of-Words (BoG) and Averaged FastText-based Sentence Embedding (AFE) on the test set.

| | BoG | AFE |
|---|---|---|
| OS C-index (95% CI) | 0.48 (0.39, 0.56) | 0.56 (0.47, 0.64) |
| DFS C-index (95% CI) | 0.56 (0.46, 0.66) | 0.59 (0.51, 0.66) |
| OS HR (95% CI) | 0.95 (0.52, 1.72) | 1.45 (0.85, 2.44) |
| Log-rank P (OS) | 0.86 | 0.17 |
| DFS HR (95% CI) | 1.44 (0.78, 2.66) | 1.54 (0.87, 2.73) |
| Log-rank P (DFS) | 0.25 | 0.14 |

95% CI: 95% confidence intervals. OS: overall survival. DFS: disease-free survival. Log-rank p: Log-rank p value. HR: Hazard Ratio.

**Supplementary Table 2.** The accuracy of masked words prediction of 5 pre-trained language models (Radiology Reports Only).

| | RobBERT | MedRoBERTa.nl | RecRoBERT | BRecRoBERT | BRec2RoBERT |
|---|---|---|---|---|---|
| Accuracy | 0.34 | 0.50 | 0.80 | 0.80 | **0.83** |
| Top 5 Accuracy | 0.49 | 0.70 | 0.90 | 0.90 | **0.91** |
| Top 10 Accuracy | 0.53 | 0.77 | 0.92 | 0.92 | **0.93** |

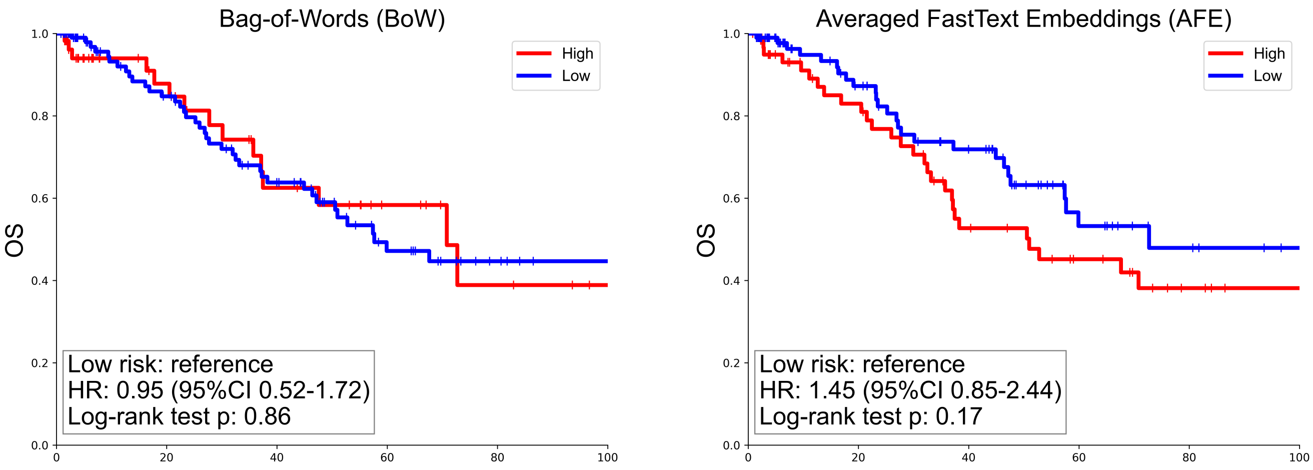


**Supplementary Figure 1.** Kaplan-Meier analysis for OS by deep learning-based risk score in the test set using BoW and AFE features.

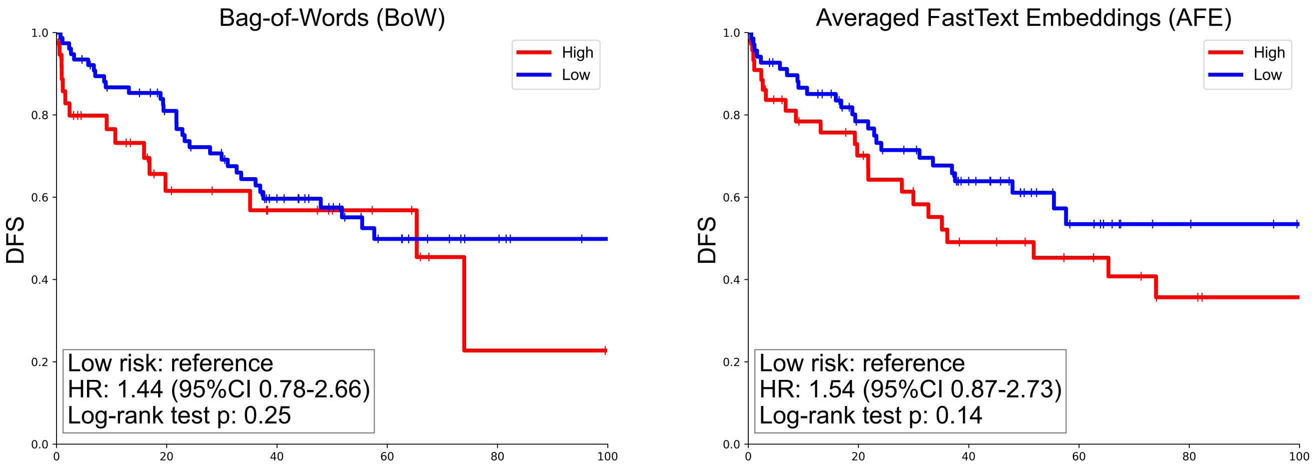


**Supplementary Figure 2.** Kaplan-Meier analysis for DFS by deep learning-based risk score in the test set using BoW and AFE features.

*Figure omitted in the preprint version.*
*This figure contains original Dutch radiology text and is therefore not displayed in accordance with medRxiv language policy.*
*The original figure is available from the corresponding author upon reasonable request.*

**Supplementary Figure 3.** Word clouds of breast cancer reports.

*Figure omitted in the preprint version.*
*This figure contains original Dutch report text and is therefore not displayed in accordance with medRxiv language policy.*
*The original figure is available from the corresponding author upon reasonable request.*

**Supplementary Figure 4.** Word clouds of Pathology and Endoscopy reports, presented separately.

*Figure omitted in the preprint version.*
*This figure contains original Dutch report text and is therefore not displayed in accordance with medRxiv language policy.*
*The original figure is available from the corresponding author upon reasonable request.*

**Supplementary Figure 5.** Scattertext analysis between rectal cancer and breast cancer reports.

*Figure omitted in the preprint version.*
*This figure contains original Dutch radiology text and is therefore not displayed in accordance with medRxiv language policy.*
*The original figure is available from the corresponding author upon reasonable request.*

**Supplementary Figure 6.** The correlation between words in a given radiological report using pre-trained ClinicalBERT.

**Supplementary Table 3.** The accuracy of masked words prediction of 5 pre-trained language models (Pathology Reports Only).

| | RobBERT | MedRoBERTa.nl | RecRoBERT | BRecRoBERT | BRec2RoBERT |
|---|---|---|---|---|---|
| Accuracy | 0.33 | 0.49 | 0.81 | 0.80 | **0.84** |
| Top 5 Accuracy | 0.51 | 0.66 | 0.91 | 0.91 | **0.93** |
| Top 10 Accuracy | 0.56 | 0.72 | 0.93 | 0.93 | **0.95** |

**Supplementary Table 4.** The accuracy of masked words prediction of 5 pre-trained language models (Endoscopy Reports Only).

| | RobBERT | MedRoBERTa.nl | RecRoBERT | BRecRoBERT | BRec2RoBERT |
|---|---|---|---|---|---|
| Accuracy | 0.33 | 0.53 | 0.88 | 0.87 | **0.90** |
| Top 5 Accuracy | 0.49 | 0.72 | 0.94 | 0.94 | **0.95** |
| Top 10 Accuracy | 0.54 | 0.77 | 0.95 | 0.95 | **0.96** |

**Supplementary Table 5.** The C-index for five pre-trained RoBERTa models in OS and DFS tasks in the interval validation.

| | RobBERT | MedRoBERTa.nl | RecRoBERT | BRecRoBERT | BRec2RoBERT |
|---|---|---|---|---|---|
| OS C-index (95% CI) | 0.61 (0.47, 0.73) | 0.59 (0.47, 0.71) | 0.55 (0.54, 0.67) | **0.68 (0.58, 0.77)** | 0.63 (0.53, 0.73) |
| DFS C-index (95% CI) | 0.60 (0.49, 0.71) | 0.61 (0.49, 0.73) | 0.75 (0.67, 0.82) | 0.75 (0.65, 0.83) | **0.83 (0.75, 0.89)** |

95% CI: 95% confidence intervals

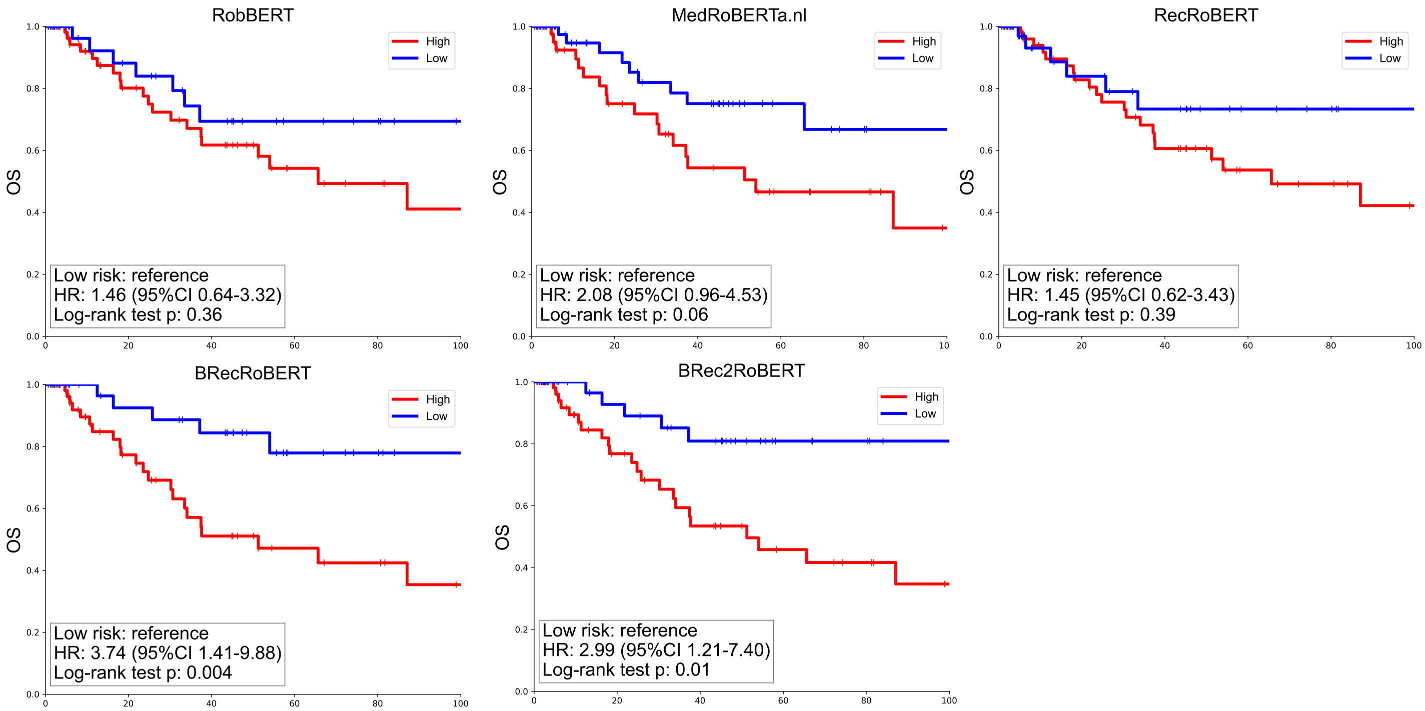


**Supplementary Figure 7.** Kaplan-Meier analysis for OS by deep learning-based risk score in the interval validation set.

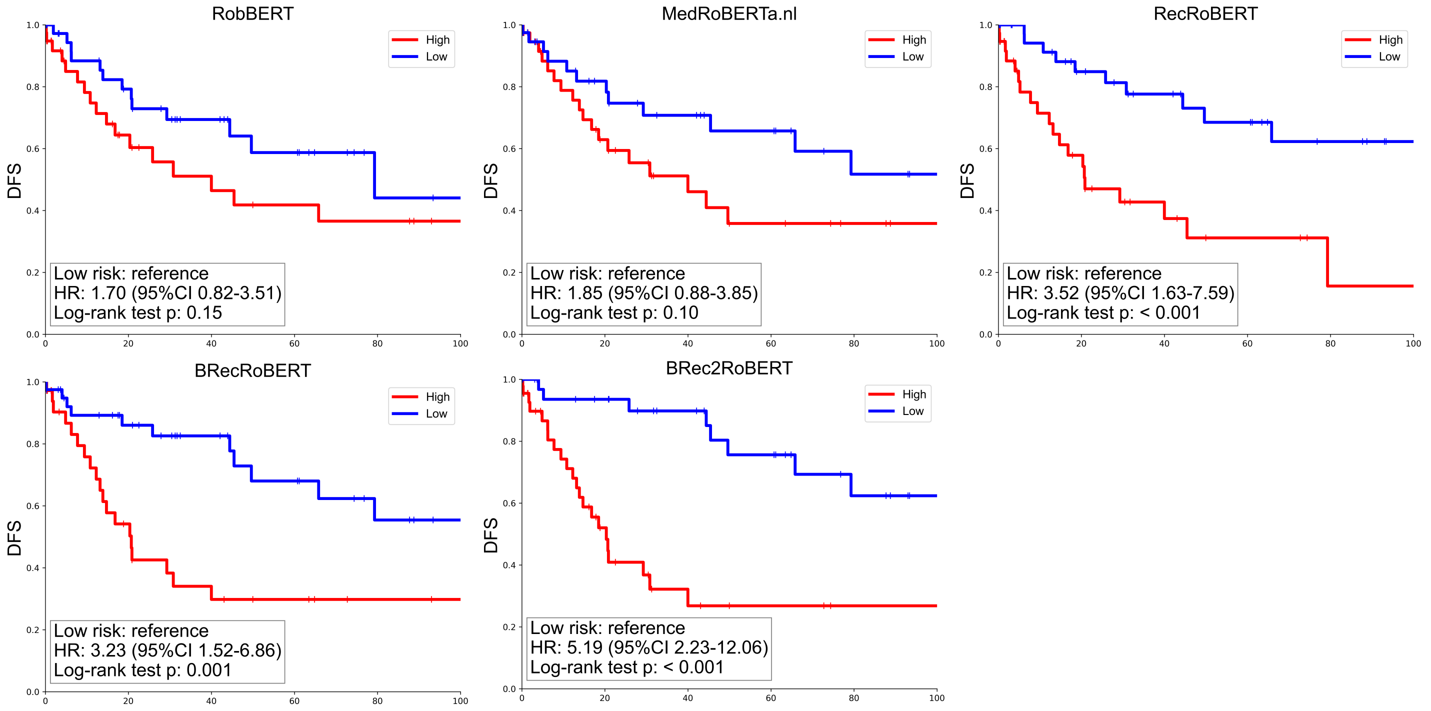


**Supplementary Figure 8.** Kaplan-Meier analysis for DFS by deep learning-based risk score in the internal validation set.

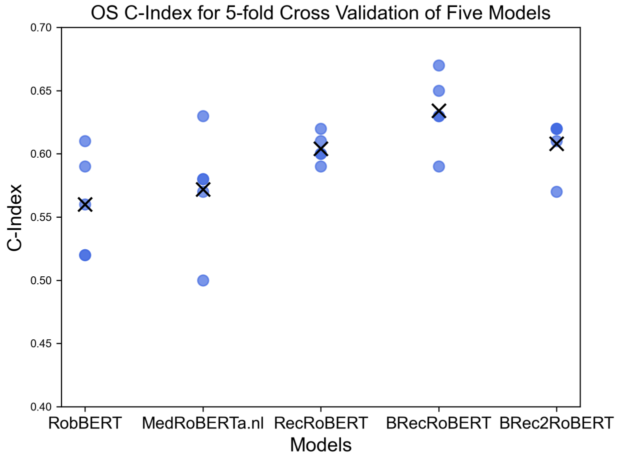


**(a)** Concordance Index (C-Index) for Overall Survival (OS) from 5-Fold Cross-Validation Across Five Models on the Test Set.

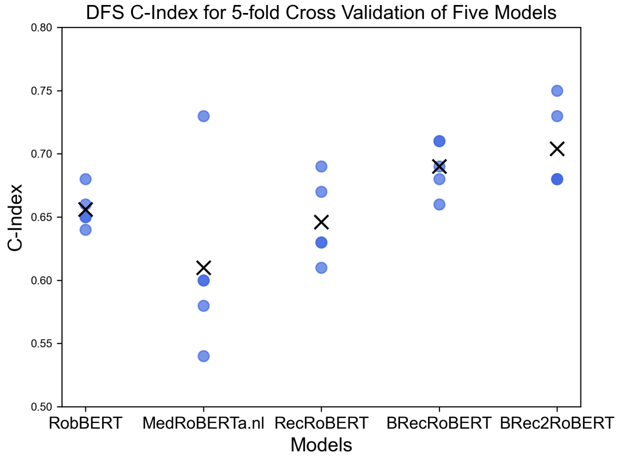


**(b)** Concordance Index (C-Index) for Disease-Free Survival (DFS) from 5-Fold Cross-Validation Across Five Models on the Test Set

**Supplementary Figure 9**

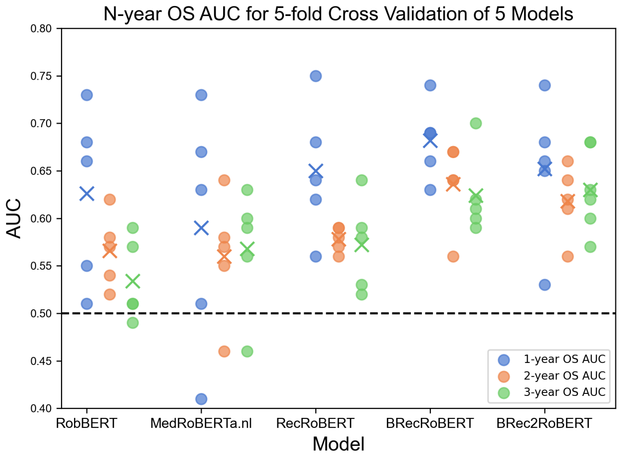


**(a)** AUC for N-year Overall Survival (OS) from 5-Fold Cross-Validation Across Five Models on the Test Set.

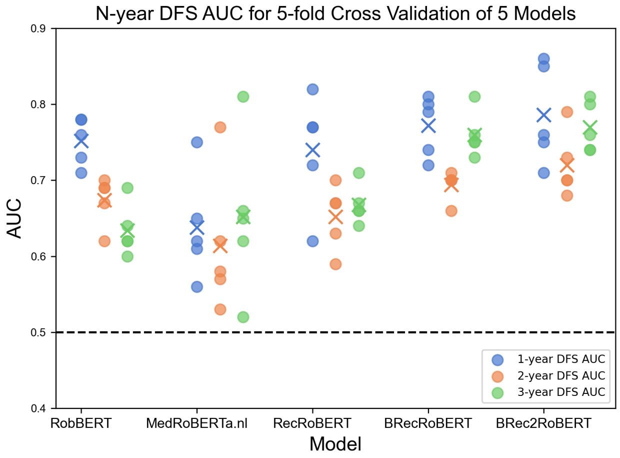


**(b)** AUC for N-year Disease-Free Survival (DFS) from 5-Fold Cross-Validation Across Five Models on the Test Set.

**Supplementary Figure 10**
